# Asymptomatic or mild symptomatic SARS-CoV-2 infection elicits durable neutralizing antibody responses in children and adolescents

**DOI:** 10.1101/2021.04.17.21255663

**Authors:** Carolina Garrido, Jillian H. Hurst, Cynthia G. Lorang, Jhoanna N. Aquino, Javier Rodriguez, Trevor S. Pfeiffer, Tulika Singh, Eleanor C. Semmes, Debra J. Lugo, Alexandre T. Rotta, Nicholas A. Turner, Thomas W. Burke, Micah T. McClain, Elizabeth A. Petzold, Sallie R. Permar, M. Anthony Moody, Christopher W. Woods, Matthew. S. Kelly, Genevieve G. Fouda

**Affiliations:** Duke Human Vaccine Institute, Duke University School of Medicine, Durham, NC; Department of Pediatrics, Division of Infectious Diseases, Duke University School of Medicine, Durham, NC; Children’s Health & Discovery Institute, Department of Pediatrics, Duke University School of Medicine, Durham, NC; Children’s Clinical Research Unit, Department of Pediatrics, Duke University School of Medicine, Durham, North Carolina, USA; Medical Scientist Training Program, Department of Molecular Genetics and Microbiology, Duke University, Durham, NC; Department of Pediatrics, Division of Pediatric Critical Care Medicine, Duke University School of Medicine, Durham, NC; Department of Medicine, Division of Infectious Diseases, Duke University School of Medicine, Durham, NC; Durham Veterans Affairs Medical Center, Durham, NC; Center for Applied Genomics and Precision Medicine, Duke University, Durham, NC; Department of Pediatrics, Weill Cornell School of Medicine, New York City, NY

**Author notes:** Corresponding Author: Genevieve G. Fouda, MD, PhD, 2 Genome Court, Durham, NC 27710. These authors contributed equally to the manuscript (Carolina Garrido, Jillian H. Hurst, Matthew. S. Kelly, Genevieve G. Fouda).

## Abstract

As SARS-CoV-2 continues to spread globally, questions have emerged regarding the strength and durability of immune responses in specific populations. In this study, we evaluated humoral immune responses in 69 children and adolescents with asymptomatic or mild symptomatic SARS-CoV-2 infection. We detected robust IgM, IgG, and IgA antibody responses to a broad array of SARS-CoV-2 antigens at the time of acute infection and 2 and 4 months after acute infection in all participants. Notably, these antibody responses were associated with virus neutralizing activity that was still detectable 4 months after acute infection in 94% of children. Moreover, antibody responses and neutralizing activity in sera from children and adolescents were comparable or superior to those observed in sera from 24 adults with mild symptomatic infection. Taken together, these findings indicate children and adolescents with mild or asymptomatic SARS-CoV-2 infection generate robust and durable humoral immune responses that are likely to protect from reinfection.

## INTRODUCTION

Severe acute respiratory syndrome coronavirus 2 (SARS-CoV-2), the etiological agent of coronavirus disease 2019 (COVID-19), has caused more than 140 million infections and nearly three million deaths globally.^1^ The effectiveness and durability of the immune responses induced by SARS-CoV-2 infection has major implications for the risk of reinfection and the establishment of herd immunity. Studies conducted among adult populations suggest that there is substantial variability in SARS-CoV-2 humoral immune responses based on patient factors and characteristics of the initial illness. In a study of more than 30,000 individuals in New York who tested positive for SARS-CoV-2 infection by PCR, more than 90% had detectable IgG antibodies to the viral spike protein within 30 days of SARS CoV-2 acute infection, irrespective of the severity of the initial illness.^2^ More recent studies have shown that antibody responses in adults are durable, with reports of detectable SARS-CoV-2-specific antibodies up to 6-8 months after acute infection.^3^ Several prior studies reported an association between the durability of antibody responses and the severity of the acute infection. For example, Long and colleagues found that 40% of the asymptomatic adults and 13% of the symptomatic adults with initial antibody responses seroreverted within two months after acute infection.^4^ Conversely, Kong and colleagues reported that subjects with mild or moderate symptoms had earlier and more robust antibody responses than subjects with severe illness during the 40 days after symptom onset.^5^ Taken together, these studies demonstrate the marked variability of immune responses within different populations and highlight a potential association between illness severity and the development of immune responses to SARS-CoV-2.

Epidemiological data from around the world indicate that children and adolescents with SARS-CoV-2 infection typically have milder illness courses than adults.^6–8^ Notably, SARS-CoV-2 infection in children and adolescents is often asymptomatic or associated with such minor symptoms that children infrequently come to medical attention.^9,10^ The varied clinical manifestations of SARS-CoV-2 infection among children and adults suggest that age may modify the host response to SARS-CoV-2, as has previously been demonstrated for several other viruses.^11–17^ To date, studies of SARS-CoV-2 immune responses in pediatric populations have focused primarily on children hospitalized for severe COVID-19 or who developed multisystem inflammatory syndrome in children (MIS-C), a potentially life-threatening inflammatory condition that can occur after SARS-CoV-2 infection.^18–22^ While these studies provide important insights into the immune responses of children and adolescents who develop these rare manifestations of SARS-CoV-2 infection, surprisingly little is known about the immune responses of the much larger population of children with asymptomatic or mild symptomatic SARS-CoV-2 infection.

In this study, we evaluated the temporal evolution of SARS-CoV-2-specific humoral immune responses in a cohort of children and adolescents with asymptomatic or mildly symptomatic SARS-CoV-2 infection. We report clinical characteristics, serum titers of antibodies specific to a panel of SARS-CoV-2 antigens, and virus neutralizing antibody responses over a period of up to 4 months after acute infection. Further, we compare the antibody responses of these children and adolescents to those of a group of adults with mild symptomatic SARS-CoV-2 infection. Knowledge of the effectiveness and durability of SARS-CoV-2-specific immune responses in children is critical to the development of pediatric vaccination strategies and approaches to mitigate SARS-CoV-2 transmission in schools and other congregate childcare settings.

## RESULTS

### Patient characteristics

We evaluated SARS-CoV-2-specific humoral immune responses in 69 children and adolescents (<21 years of age) who participated in a study of acute SARS-CoV-2 infection.^6^ Clinical data and sera were collected from these participants during the acute phase of infection and approximately 2 and 4 months after acute infection. Median [interquartile range (IQR)] age was 11.5 (5.2, 16.5) years, and 51% were female (**Table 1**). The most common chronic medical conditions were obesity (29%) and asthma (6%). Fifty-five (80%) participants reported one or more symptoms associated with acute SARS-CoV-2 infection with a median (IQR) symptom duration of 3 (2, 7) days. The most common symptoms reported were fever (49%), cough (36%), and headache (29%). The remaining 14 (20%) participants reported no symptoms associated with acute infection (2 weeks prior to SARS-CoV-2 diagnosis and up to 28 days after study enrollment) and were classified as asymptomatic. Sera from acute infection was collected at a median (IQR) of 9 (6, 12) days from symptom onset or SARS-CoV-2 diagnosis in asymptomatic subjects. Sera from 2 months and 4 months after acute infection were collected at a median (IQR) of 57 (50, 71) days and 109 (101, 120) days from symptom onset or SARS-CoV-2 diagnosis, respectively. To compare humoral immune responses across the full spectrum of age, we similarly evaluated humoral immune responses to SARS-CoV-2 in 24 adults (21 years of age or older). Median (IQR) age was 43 (31, 57) years, and 58% were female (**Table 1**). All adult participants reported one or more symptoms associated with acute SARS-CoV-2 infection. No pediatric or adult subjects required hospitalization for SARS-CoV-2 infection.

**Table 1.** Demographic and clinical features of the study cohort by age

|  | **0-5 years**<br>**(n=20)** |  | **6-13 years**<br>**(n=26)** |  | **14-20 years**<br>**(n=23)** |  | **21-70 years**<br>**(n=24)** |  |
| --- | --- | --- | --- | --- | --- | --- | --- | --- |
| Median (IQR) age, | 2.1 | (0.9, 4.0) | 9.5 | (8.1, | 17.1 | (16.5, | 43.4 | (31.2, 56.8) |
| Sex |  |  |  |  |  |  |  |  |
| Female | 9 | 45% | 11 | 42% | 15 | 65% | 14 | 58% |
| Male | 11 | 55% | 15 | 58% | 8 | 35% | 10 | 42% |
| Illness severity |  |  |  |  |  |  |  |  |
| Asymptomatic | 2 | 10% | 8 | 31% | 4 | 17% | 0 | 0% |
| Mild symptomatic | 18 | 90% | 18 | 69% | 19 | 83% | 24 | 100% |
| Serum samples |  |  |  |  |  |  |  |  |
| 0 months | 20 | 100% | 26 | 100% | 23 | 100% | 22 | 92% |
| 2 months | 17 | 85% | 22 | 85% | 17 | 74% | 19 | 79% |
| 4 months | 12 | 60% | 19 | 73% | 19 | 83% | 14 | 58% |

IQR, interquartile range

### SARS-CoV-2-specific antibodies in sera from children and adolescents

We used Luminex-based binding antibody multiplex assays to measure SARS CoV-2–specific antibodies targeting whole spike, subunit 1 (S1), receptor binding domain (RBD), N-terminal domain (NTD), subunit 2 (S2), nucleocapsid (NC), and membrane (M) proteins in sera from children and adolescents during acute infection (n=69), 2 months after acute infection (n=56), and 4 months after acute infection (n=50). At the time of acute infection, all children and adolescents had detectable levels of IgM and IgG antibodies to one or more SARS-CoV-2 antigens, and 68 (99%) had a detectable IgA response to one or more SARS-CoV-2 antigens. Levels of SARS-CoV-2-specific IgM and IgA antibodies were highest during acute infection and declined at 2 and 4 months after acute infection, while levels of IgG antibodies generally increased from acute infection to 2 months after infection before decreasing between 2 and 4 months after acute infection (**Figure 1, Supplemental Figure 1**, and **Supplemental Table 1**). We also measured levels of IgG subclasses to SARS-CoV-2 antigens at these same time points (**Supplemental Figure 2a**). Between acute infection and 2 months after acute infection, we observed significant increases in the ratios of IgG1:IgG3 antibodies to spike (p=0.04), S1 (p<0.0001), RBD (p<0.0001), S2 (p=0.03), and NC (p<0.0001); these differences persisted comparing acute infection to 4 months after acute infection for these same antigens (spike: p<0.0001; S1: p<0.0001; RBD: p<0.0001; S2: p<0.0001; NC: p<0.0001). No differences in the ratios of IgG1:IgG3 antibodies to NTD or M were observed over time (**Supplemental Figure 2b**).

**Figure 1.**
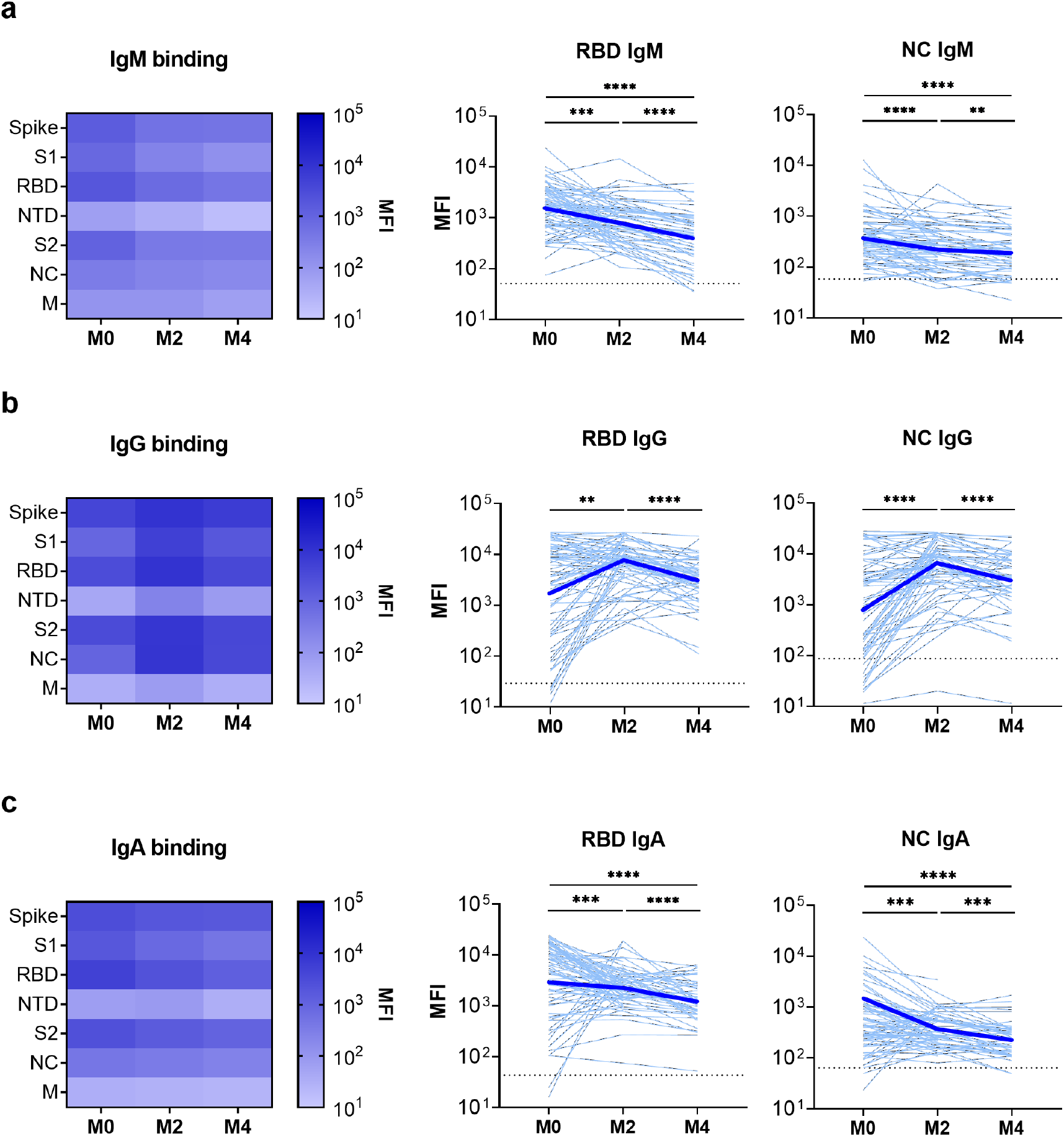
SARS-CoV-2-specific binding antibodies in children and adolescents after acute infection. Specific binding to SARS-CoV-2 antigens was measured by Luminex-based multiplex assays for IgM (**a**), IgG (**b**), and IgA (**c**) antibodies to the whole spike, subunit 1 (S1), receptor binding domain (RBD), N-terminal domain (NTD), subunit 2 (S2), nucleocapsid (NC), and membrane (M) proteins. Binding is expressed as mean fluorescence intensity (MFI) of sera at the time of acute infection (M0) and 2 months (M2) and 4 months (M4) after acute infection. Heatmaps show binding to all analyzed SARS-CoV-2 antigens, with darker colors corresponding to higher binding. Line graphs depict RBD- and NC-specific binding of sera from individual participants (light blue lines) and the geometric mean of all individuals (thick blue lines). Dotted lines indicate assay positivity thresholds and were calculated as the mean MFI plus three standard deviations using sera from 10 SARS-CoV-2-uninfected individuals. Comparisons of samples from individuals across time points were made using Wilcoxon signed-rank tests. * p<0.05; ** p<0.01; *** p<0.005; **** p<0.0001

**Figure 2.**
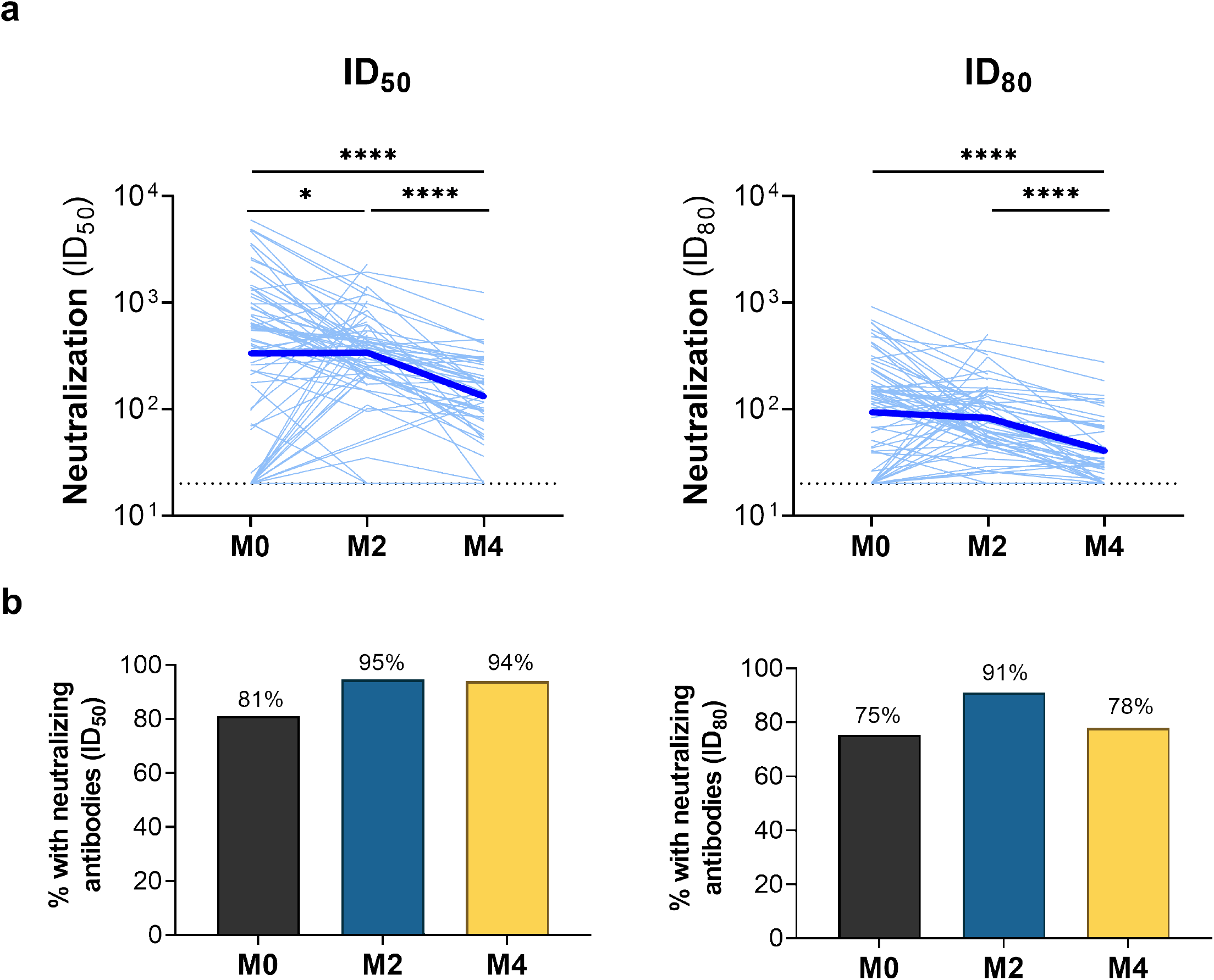
SARS-CoV-2 neutralizing antibodies in sera from children and adolescents. Antibody-mediated neutralization activity was measured using a pseudovirus (614G) assay and is presented as 50% inhibitory dilution (ID50) or 80% inhibitory dilution (ID80). (**a**) Neutralization activity in sera from specific individuals (light blue lines) and the geometric mean of all individuals (thick blue lines) is shown at the time of acute infection (M0) and 2 months (M2) and 4 months (M4) after acute infection. Dotted lines indicate assay positivity thresholds. Comparisons of samples from individuals across time points were made using Wilcoxon signed-rank tests. (**b**) Proportion of children with detectable neutralizing antibodies at ID_50_ (left) or ID_80_ (right) at specific time points after acute infection. * p<0.05; ** p<0.01; *** p<0.005; **** p<0.0001

### SARS-CoV-2-infected children and adolescents generate durable neutralizing antibody responses

We next measured neutralizing activity of sera from these children and adolescents using a luciferase-based SARS-CoV-2 pseudovirus (614G) assay, as previously described.^23^ Neutralizing antibodies at a 50% inhibitory dilution (ID_50_) were detected in 56 of 69 (81%) serum samples collected during acute infection, 53 of 56 (95%) samples collected 2 months after acute infection, and 47 of 50 (94%) of samples collected 4 months after acute infection (**Figure 2**). Acute infection samples for which there was no appreciable neutralizing activity were collected earlier after symptom onset or SARS-CoV-2 diagnosis than acute infection samples with detectable neutralizing antibodies [median (IQR) of 7 (5, 8) vs. 9 (7, 13) days; p=0.01]. Between acute infection and 2 months after acute infection, levels of neutralizing antibodies declined in 31 (55%) children, increased in 24 (43%) children, were stable in one (2%) child (median ID_50_: 577 vs. 379; p=0.04). Neutralizing antibodies declined in 31 of 37 (84%) children with paired samples at 2 and 4 months after acute infection (median ID_50_: 369 vs. 174; p<0.0001). As anticipated, neutralizing activity measured in this pseudovirus assay was most strongly correlated with levels of antibodies to spike, S1, and RBD (**Supplemental Figure 3**).

**Figure 3.**
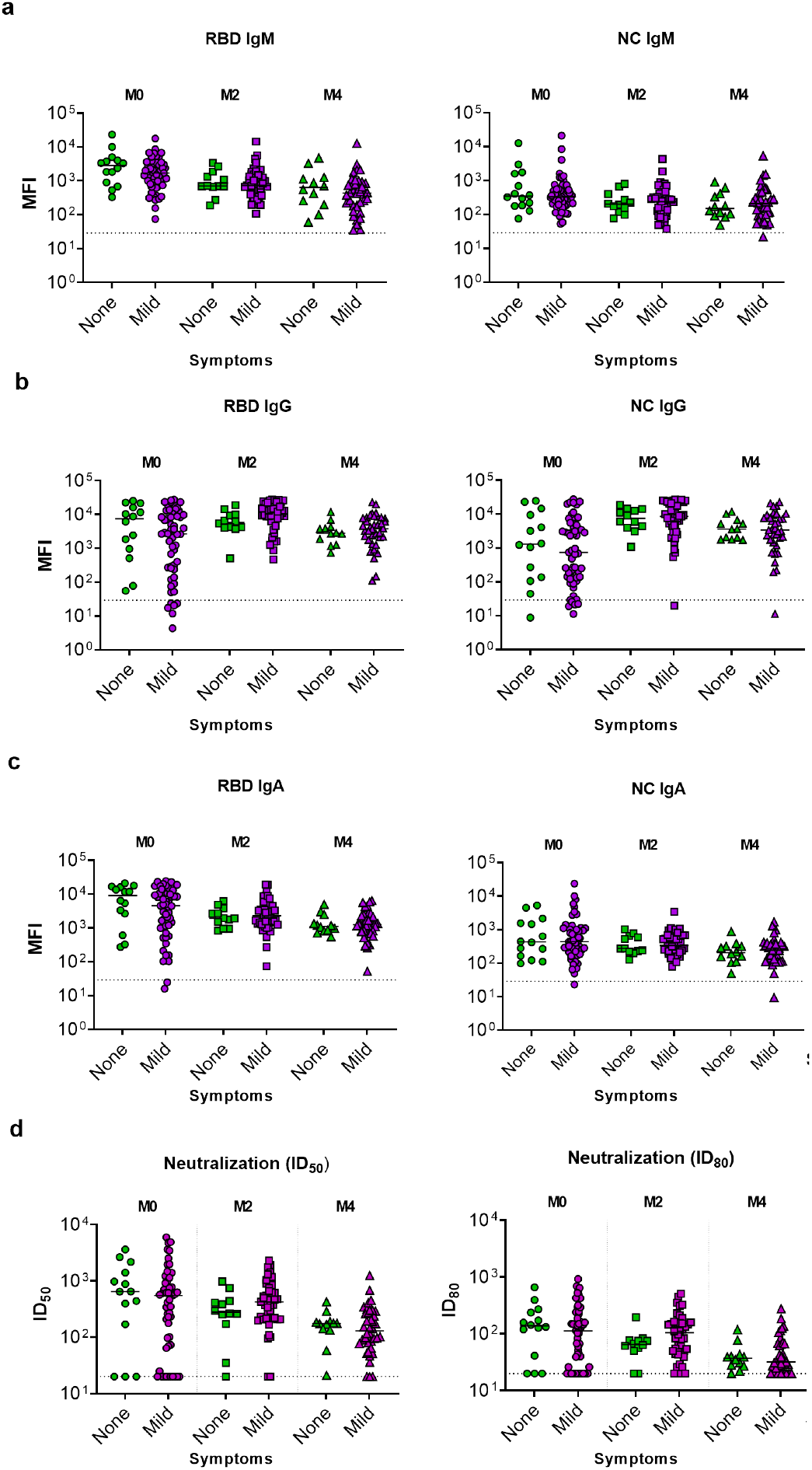
SARS-CoV-2-specific antibodies and neutralizing activity in sera from children and adolescents with asymptomatic compared with mild symptomatic infection. Levels of IgM (**a**), IgG (**b**), and IgA (**c**) antibodies to SARS-CoV-2 receptor binding domain (RBD) and nucleocapsid (NC) proteins were measured by Luminex-based multiplex assays and are expressed as mean fluorescence intensity (MFI). Binding is shown at the time of acute infection (M0) and 2 months (M2) and 4 months (M4) after acute infection. Dotted lines for binding assays correspond to the mean MFI plus 3 standard deviations in sera from 10 SARS-CoV-2-uninfected individuals. (**d**) Antibody-mediated neutralization activity was measured using a pseudovirus (614G) assay and is presented as 50% inhibitory dilution (ID50) or 80% inhibitory dilution (ID80). No significant differences in levels of antibodies to specific SARS-CoV-2 antigens or neutralizing activity were seen at any time point among children with asymptomatic versus mild symptomatic infection (Wilcoxon rank-sum tests).

### SARS-CoV-2-specific antibody responses are similar in asymptomatic and mildly symptomatic children

Given that prior studies in adults suggested that humoral immune responses to SARS-CoV-2 infection may differ based on symptom severity,^5,24–29^ we compared antibody responses among children and adolescents with asymptomatic and mild symptomatic SARS-CoV-2 infection. We observed no significant differences in the levels of IgM, IgG, or IgA antibodies specific to any SARS-CoV-2 antigen during acute infection or at 2 or 4 months after acute infection in asymptomatic and mildly symptomatic children (**Figure 3a-c**). Similarly, the degree of pseudovirus neutralization activity in sera did not differ based on the presence of symptoms at any of these time points (**Figure 3d**).

### Comparisons of SARS-CoV-2 antibody responses in children, adolescents, and adults

We next compared the SARS-CoV-2 humoral immune responses of children and adolescents to those of adults. IgM and IgG antibodies to at least one SARS-CoV-2 antigen were detected in all adults at each time point; IgA antibodies were found in 22 of 22 (100%) samples collected during acute infection, 18 of 19 (95%) samples collected at 2 months after acute infection, and 11 of 14 (79%) samples collected at 4 months after acute infection (**Supplemental Figure 4** and **Supplemental Table 2**). In order to evaluate for age-related differences in humoral immune responses to SARS-CoV-2, we compared levels of SARS-CoV-2-specific antibodies in children, adolescents, and adults at each time point. RBD-specific IgM titers were largely similar across age groups during acute infection and 2 months after acute infection (**Figure 4a**); however, levels of RBD IgM antibodies declined more quickly in children 0-5 years of age and differed from all other age groups at 4 months after acute infection. NC-specific IgM titers tended to be lower in children 0-5 years of age or 6-13 years of age at all time points. Similar trends were observed by age for IgM antibodies to other SARS-CoV-2 antigens with the exception of M protein, as adults had significantly lower levels of M-specific IgM antibodies than children or adolescents at all time points (**Supplemental Figure 5a**). Levels of RBD-specific IgG were generally similar across age groups during acute infection (**Figure 4b**), but all children, regardless of age group, had higher levels of RBD IgG than adults at 2 months [median (IQR) MFI: 9360 (4520, 14451) vs. 2028 (965, 4022); p=0.0001] and 4 months after acute infection [median (IQR) MFI: 3494 (1941, 6721) vs. 1043 (414, 2379); p=0.02]. Similar differences by age across these time points were observed for IgG antibodies specific to NC (**Figure 4b**) and other SARS-CoV-2 antigens (**Supplemental Figure 5b**). Levels of IgA antibodies to spike, S2, and NC were generally lower among children 0-5 years or 6-13 years of age than amongst older age groups at all time points (**Supplemental Figure 5c**).

**Figure 4.**
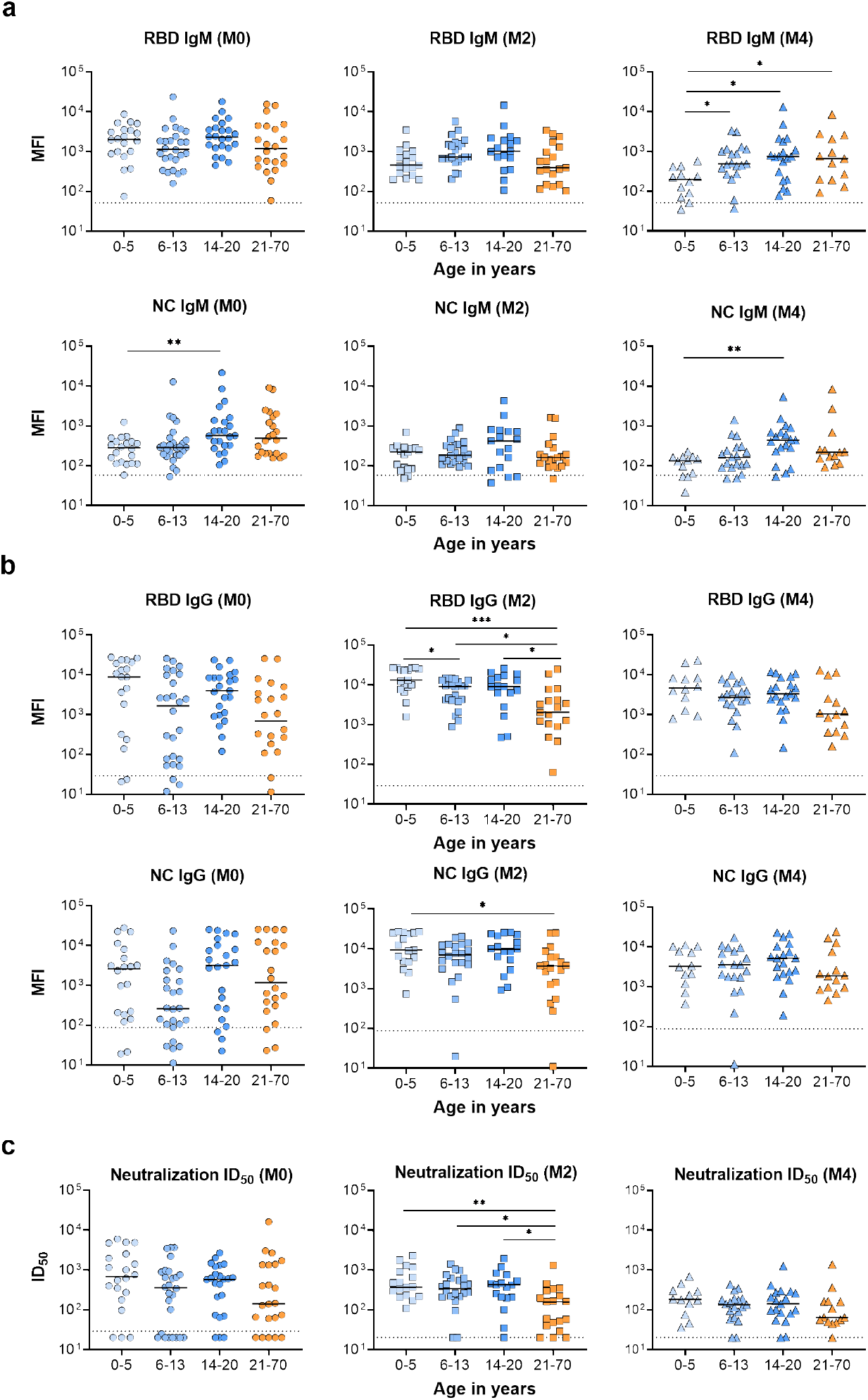
SARS-CoV-2-specific antibodies and neutralizing activity in sera from children and adults by age. Levels of IgM (**a**) and IgG (**b**) antibodies to SARS-CoV-2 receptor binding domain (RBD) and nucleocapsid (NC) proteins were measured by Luminex-based multiplex assays and are expressed as mean fluorescence intensity (MFI). Dotted lines for binding assays correspond to the mean MFI plus three standard deviations in sera from 10 SARS-CoV-2-uninfected individuals. (**c**) Antibody-mediated neutralization activity was measured in a pseudovirus (614G) assay and is presented as 50% inhibitory dilution (ID50). Dotted lines for neutralization assays correspond to the assay threshold. Comparisons of antibody measures by age category were performed at acute infection (M0) and 2 months (M2) and 4 months (M4) after acute infection using Wilcoxon rank-sum tests and adjusting for multiple comparisons using the Benjamini-Hochberg procedure. * p<0.05, ** p<0.01, *** p<0.005

Virus neutralizing activity at ID50 was detected in 17 of 22 (77%) serum samples collected from adult participants during acute infection, 16 of 19 (84%) samples collected at 2 months after acute infection, and 13 of 14 (93%) samples collected at 4 months after acute infection (**Supplemental Figure 6**). As observed among children and adolescents, acute infection samples for which there was no appreciable neutralizing activity were collected earlier after symptom onset than acute infection samples with detectable neutralizing antibodies [median (IQR) of 4 (3, 8) vs. 16 (12, 20) days; p=0.004]. Neutralizing activity in sera was similar across age groups at the time of acute infection. However, paralleling the trends observed for SARS-CoV-2-specific IgG antibodies, children and adolescents tended to have higher neutralizing activity than adults at 2 months [median (IQR) ID50: 379 (214, 634) vs. 161 (44, 262); p=0.0003] and 4 months after acute infection [median (IQR) ID50: 148 (81, 254) vs. 64 (52, 162); p=0.10], although these differences were not statistically significant at 4 months (**Figure 4c**).

## DISCUSSION

In this study, we describe the durability and functionality of the humoral immune responses of children and adolescents with asymptomatic or mild symptomatic SARS-CoV-2 infection. SARS-CoV-2 infection elicited robust neutralizing antibody responses during acute infection and up to 4 months after acute infection in children and adolescents. Moreover, these antibody responses did not differ among children with mild symptomatic infection compared to those with asymptomatic infection, suggesting that effective humoral responses are elicited regardless of the severity of the acute infection. Notably, we found that the SARS-CoV-2-specific antibody responses of children and adolescents were generally more robust and durable than those of adults with mild symptomatic infection.

Several prior studies evaluated the prevalence and durability of SARS-CoV-2 infection-induced humoral immune responses in adult populations. Studies conducted early in the COVID-19 pandemic reported that seroconversion occurs in approximately 90-95% of SARS-CoV-2-infected adults, with SARS-CoV-2-specific IgM, IgG, and IgA antibodies detectable in most individuals within two weeks of symptom onset.^2,30–35^ Similarly, we found that 100% of children in our study exhibited IgM and IgG responses and 99% exhibited IgA responses within this time period. Multiple reports have demonstrated that SARS-CoV-2-specific antibody responses among adults are durable, with a recent study reporting that IgG to the SARS-CoV-2 spike protein is detectable at least six months after infection in more than 90% of adults.^3^ Neutralizing antibody responses, which are believed to correlate most closely with protection from reinfection,^36,37^ are reportedly stable for at least 3 months after infection in adults.^38,39^ Similarly, seroconversion occurred in all children and adolescents evaluated in our study. Neutralizing activity was more likely to be detectable in sera collected from both child and adults participants who were slightly further into the course of their acute infection, indicating a slight lag in the development of an effective neutralizing antibody response. Importantly, SARS-CoV-2-specific humoral immune and virus neutralizing responses were detectable in the vast majority of subjects 4 months after acute infection, demonstrating that neutralizing antibody responses in children are similar in duration to those in adults.

To date, most studies of the immune responses of children to SARS-CoV-2 have been cross-sectional, have focused on patients hospitalized for severe COVID-19 or MIS-C, or have assessed immunity only during acute infection.^22^ Weisberg and colleagues evaluated SARS-CoV-2-specific antibody levels and neutralizing activity in non-hospitalized adults with COVID-19, hospitalized adults with COVID-19 acute respiratory distress syndrome, children with MIS-C, and SARS-CoV-2-infected children, half of whom were asymptomatic.^18^ They found no significant differences in antibody levels or neutralizing activity between the two groups of children; however, both groups of adults exhibited higher SARS-CoV-2-specific antibody levels and neutralizing activity than either group of children.^18^ Similarly, Pierce and colleagues found that children who were hospitalized with COVID-19 or MIS-C had lower levels of neutralizing activity than hospitalized adults.^21^ In our study, we found that SARS-CoV-2-specific antibody levels and virus neutralizing activity among children and adolescents with asymptomatic or mild symptomatic SARS-CoV-2 infection were generally similar to those of adults with mild symptomatic infection. Importantly, levels of SARS-CoV-2-specific IgG and serum neutralizing activity were similar at the time of acute infection but generally higher in children and adolescents than adults at 2 and 4 months after acute infection, suggesting that SARS-CoV-2-specific IgG responses may decline more slowly in children and adolescents. In contrast, we observed higher levels of SARS-CoV-2-specific IgM and IgA antibodies with increasing age, particularly 4 months after acute infection, suggesting that young children generate less robust and durable responses for these SARS-CoV-2-specific antibody isotypes. Overall, our findings indicate that children and adolescents have a similar degree of protective immunity as adults after mild or asymptomatic SARS-CoV-2 infection. Given similarities in the response to natural infection in children and adults, it is likely that vaccination against SARS-CoV-2 will also elicit a similar degree of protection across the full spectrum of age, as has recently been reported for the Pfizer-BioNTech vaccine in children 12-15 years of age.^40^ Though we cannot directly compare our results to the neutralizing antibody titers reported in vaccine trial studies, that younger age may be associated with greater neutralizing antibody responses. Together with our results, these clinical trial results demonstrated the ability of children to elicit robust SARS CoV-2-specific antibody responses. Future studies will need to directly evaluate associations between age and SARS-CoV-2 vaccine responses.

There are conflicting data regarding the impact of illness severity on immune responses to SARS-CoV-2 infection. Lau and colleagues found that the neutralizing activity of sera from 195 adults and children with prior SARS-CoV-2 infection was correlated with disease severity, as classified by presence of symptoms and need for supplemental oxygen.^24^ Similarly, Dogan and colleagues reported a positive correlation between level of care required and elevated SARS-CoV-2-specific antibody levels and neutralization activity in a study of 115 adult participants.^25^ Supporting this finding, several other groups reported lower humoral immune responses in patients with mild or asymptomatic disease, including a complete lack of detectable levels of circulating SARS-CoV-2-specific immunoglobulins in some cases.^26–29^ In contrast, we observed similar levels of SARS-CoV-2-specific IgM, IgG, and IgA antibodies and neutralizing activity up to 4 months after acute infection in asymptomatic and mildly symptomatic children. Given that severe illness is uncommon in SARS-CoV-2-infected children and adolescents, our findings indicate that the vast majority of children and adolescents with SARS-CoV-2 infection can be anticipated to generate robust and durable immune responses to the virus.

Our study has several limitations. First, we focused on pediatric and adult populations with asymptomatic and mild infections, but did not include participants with severe COVID-19 or children with MIS-C; thus, our findings are generalizable only to individuals with asymptomatic or mild symptomatic SARS-CoV-2 infection. Second, we did not evaluate cellular immunity, which is likely required for long-term immunological memory.^3^ Third, while several studies in adults have measured antibody responses out to 8 months after acute infection, we currently only have data for children and adolescents up to 4 months after infection. Finally, additional studies will be needed to assess the impact of emerging SARS-CoV-2 variants on viral-specific immune responses in children and adolescents and define if immune responses induced by prior infections are able to protect against these variants.

In conclusion, we found that children and adolescents with asymptomatic or mild symptomatic SARS-CoV-2 infection mount broad, effective, and durable antibody responses that exhibit robust viral neutralizing activity at least 4 months after acute infection. Notably, these responses were largely similar or superior to those observed in adults with symptomatic SARS-CoV-2 infection who did not require hospitalization. Our findings suggest that children and adolescents develop effective humoral immune responses irrespective of illness severity that are likely to provide protection against reinfection, thereby contributing to the establishment of herd immunity.

## ONLINE METHODS

### Study Design

The Duke Biospecimens from RespirAtory Virus-Exposed Kids (BRAVE Kids) study is a prospective cohort study of children and adolescents (<21 years of age) with confirmed SARS-CoV-2 infection or close contact with an individual with confirmed SARS-CoV-2 infection.^6^ This study is being conducted within the Duke University Health System (DUHS) in Raleigh-Durham, North Carolina. DUHS is a large, integrated health system consisting of three hospitals and over 100 outpatient clinics. This study was approved by the DUHS Institutional Review Board. Informed consent was obtained from all study participants or their legal guardians, with written approval obtained using an electronic consent document.

### Study Procedures

The analyses presented herein were limited to participants with SARS-CoV-2 infection diagnosed by PCR between May 1, 2020 and July 31, 2020. SARS-CoV-2 was detected from nasopharyngeal or nasal swabs through PCR testing performed for clinical care or through a quantitative real-time PCR assay, as previously described.^6^ We collected exposure, sociodemographic, and clinical data at the time of enrollment through review of electronic medical records and a directed caregiver questionnaire conducted by telephone. In addition, we conducted a home visit to collect whole blood from participants via venipuncture and to obtain other biospecimens. Serum was isolated from whole blood via centrifugation and frozen to -80°C prior to analysis. We conducted follow-up study visits at home or at a research clinic site approximately 2 and 4 months after acute infection.

### Adult Study Participants

We obtained previously collected sera from subjects enrolled in the Duke University Molecular and Epidemiological Study of Suspected Infection (MESSI)^42^. Participants in this prospective cohort are identified via community-enrollment, DUHS, or the Durham Veterans Affairs Health System (DVAHS) as having SARS-CoV-2 infection by PCR testing performed at either the North Carolina State Laboratory of Public Health or through the clinical laboratories at either DUHS or DVAHS. Participants were sampled between March and December 2020. This study was approved by the DUHS Institutional Review Board. Informed consent was obtained from all study participants, with written approval obtained using an electronic consent document. Mild disease was defined as any PCR-confirmed infection that did not require hospitalization.

### Measurement of serum SARS-CoV-2-specific antibodies

We measured serum antibodies using a customized binding antibody multiplex assay to the following SARS-CoV-2 antigens: whole spike (S), subunit 1 (S1), subunit 2 (S2), receptor binding domain (RBD), N-terminal domain (NTD), nucleocapsid (NC), and membrane (M) proteins (all from Sinobiological except from M, from MyBiosource). SARS-CoV-2 antigens were covalently coupled to magnetic fluorescent beads (MagPlex biospheres, Luminex). Unconjugated (blank) beads were included to monitor non-specific binding. After a pilot assay to identify the optimal serum dilution, antigen-coupled beads were incubated with a 1:400 serum dilution for measurement of IgG and IgG subclasses and 1:100 serum dilution for measurement of IgA and IgM. Antibody binding to the bead-coupled antigens was then detected with phycoerythrin (PE)-conjugated mouse anti-human IgG, IgM or IgA (Southern Biotech) at 2 μg/ml, using a Bio-Plex 200 instrument (Bio-Rad Laboratories), which rendered a mean fluorescent intensity (MFI) for each sample. For measurement of SARS CoV-2-specific IgG1 and IgG3, a biotinylated mouse anti-human IgG-1 or IgG-3 followed by PE-conjugated streptavidin was used for detection. Sera from ten individuals collected before the COVID-19 pandemic (2013-2014) was used to define the assay positivity threshold for each antigen (mean MFI plus 3 standard deviations). A prescreened pooled serum sample of two unrelated SARS-CoV-2 infected donors was used as positive control in all assays to ensure reproducibility between assays and to ensure detection of antibodies against all antigens tested. Criteria for accepting results included ≤20% coefficient of variation of the two duplicates with a bead count of ≥100 for each sample.

### SARS-CoV-2 Pseudovirus Neutralization Assays

SARS-CoV-2 neutralization was measured with spike-pseudotyped viruses in HEK-293T-hACE2 cells as a function of reduction in luciferase (Luc) reporter activity. We used an *env*-deficient HIV-based lentiviral system to produce viral particles pseudotyped with SARS-CoV-2 spike. An expression plasmid encoding the codon-optimized full-length spike protein of the Wuhan-1 strain (VRC7480) was provided by the Vaccine Research Center at the National Institutes of Health. The D614G amino acid change was introduced into VRC7480 by site-directed mutagenesis using the QuikChange Lightning Site-Directed Mutagenesis Kit (Agilent Technologies) and confirmed by full-length spike gene sequencing. Pseudovirions were produced in HEK 293T/17 cells (ATCC) by using FuGene 6 (Promega) and co-transfecting spike plasmid, lentiviral backbone plasmid (pCMV ΔR8.2), a firefly luciferase reporter gene plasmid (pHR’ CMV Luc) [1], and a plasmid containing TMPRSS2 (required for cell entry). TCID50 assays were performed on thawed pseudovirus aliquots using HEK-293T-hACE2 cells (BEI) to determine the infectious dose for neutralization assays.

For neutralization, serum samples were heat-inactivated for 30 minutes at 56°C prior to testing. Samples were serially diluted 5-fold to 8 points in duplicate and incubated with a dose of pre-titrated pseudovirus for 1-1.5hr at 37°C in 96-well flat-bottom poly-L-lysine-coated culture plates (Corning Biocoat). HEK-293T-hACE2 cells were suspended using TrypLE Select Enzyme solution (Thermo Fisher Scientific) and immediately added to all wells (10,000 cells in 100 µL of growth medium per well). One set of 8 control wells received cells + virus (virus control) and another set of 8 wells received cells only (background control). After 72 hrs of incubation, medium was removed and 30 µL of Promega 1X lysis buffer was added. After a 10-minute incubation at room temperature, 100 µl of Bright-Glo luciferase reagent (Promega) was added to all wells. After 2 minutes, 110 µl of the cell lysate was transferred to a black/white plate. Luminescence was measured using a PerkinElmer Life Sciences, Model Victor3 luminometer. Neutralization titers represent the serum dilution at which relative luminescence units (RLU) were reduced by either 50% (ID50) or 80% (ID80) compared to virus control wells after subtraction of background RLUs.

### Data Analysis

We described characteristics of the study population using frequencies and percentages for categorical variables and medians and interquartile ranges (IQR) for continuous variables. We used chi-squared or Fisher’s exact tests for comparisons of categorical variables. To compare humoral immune responses at specific time points based on the presence of symptoms and across age categories, we used Wilcoxon rank-sum tests or ANOVA based on Gaussian distribution of a variable. To account for repeated measurements from subjects, we used Wilcoxon signed-rank tests to compare humoral immune responses in paired serum samples collected from individuals across specific time points. We adjusted for multiple comparisons using the Benjamini-Hochberg procedure.^43^ Study data were managed using REDCap electronic data capture tools hosted at Duke University.^44^ Analyses were performed using GraphPad Prism (GraphPad Software, Inc., CA, US) and R version 4.0.3.^45^

## Data Availability

Data available upon request to the authors.

## Acknowledgments

We offer sincere gratitude to the children and families who participated in this research. We thank Dr. David Montefiori for providing the primers used in the SARS-CoV-2 pseudovirus neutralization assay.

## Declarations

TWB is a consultant for Predigene. CWW reports advisory board fees from Roche Molecular Sciences, non-financial support from bioMérieux and Becton Dickinson, a research collaboration with Biofire, and is co-founder of Predigen. MSK reports advisory board feeds from Adagio Therapeutics, Inc. All other authors have no competing interests to declare.

---

**Supplemental Figure 1.**
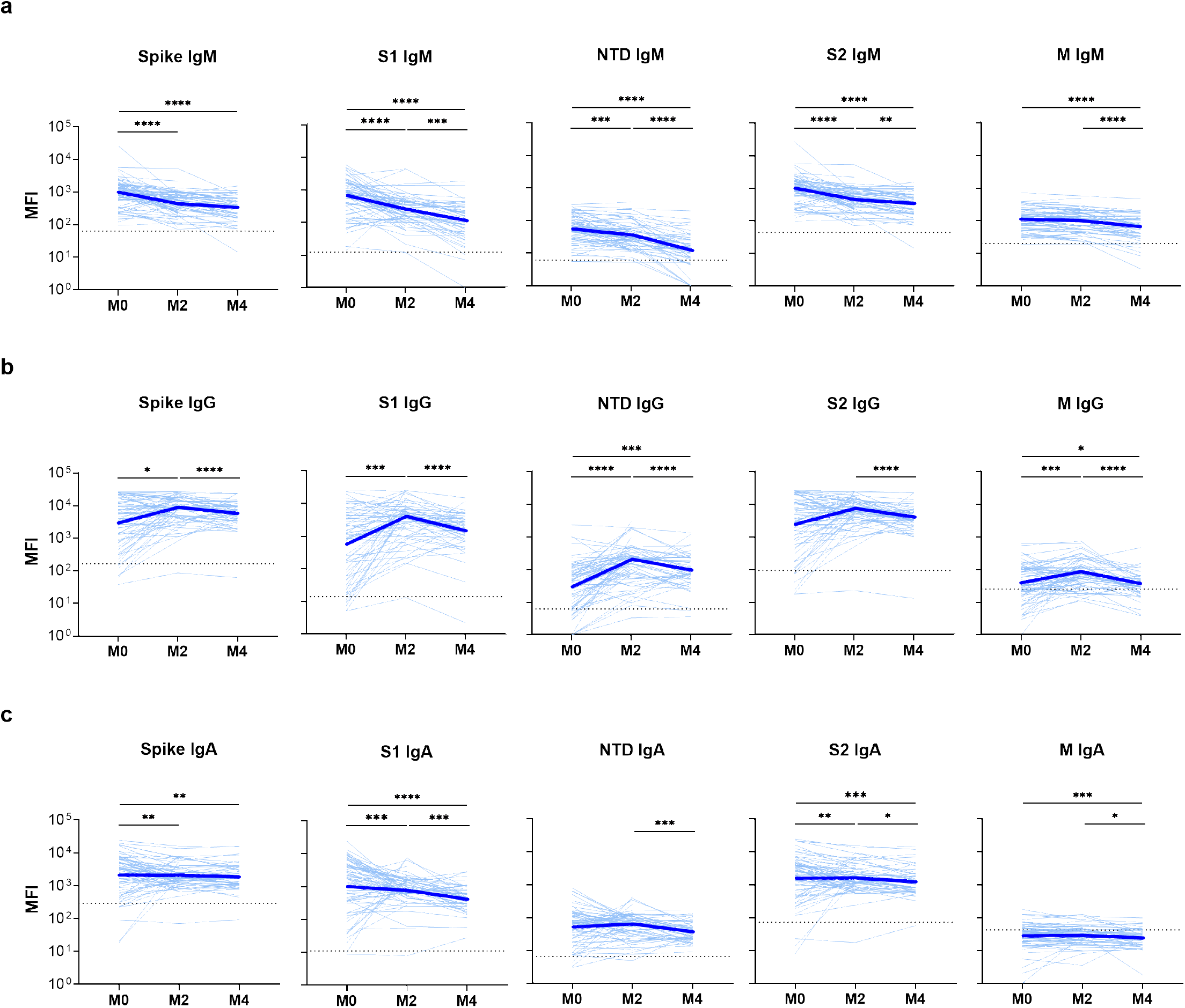
SARS-CoV-2-specific binding antibodies in sera from children and adolescents. Binding to specific SARS-CoV-2 antigens including spike, subunit 1 (S1), N-terminal domain (NTD), subunit 2 (S2), and membrane (M) proteins was measured by a Luminex-based multiplex assay for IgM (**A**), IgG (**B**), and IgA (**C**). Mean fluorescence intensity (MFI) of sera from specific individuals (light blue lines) and the geometric mean in all individuals (thick blue lines) are shown at the time of acute infection (M0) and 2 months (M2) and 4 months (M4) after acute infection. The dotted lines indicate assay thresholds corresponding to the mean MFI plus three standard deviations in sera from 10 SARS-CoV-2-uninfected individuals. Levels of specific antibodies were compared across time points using Wilcoxon signed-rank tests. * p<0.05; ** p<0.01; *** p<0.005; **** p<0.0001

**Supplemental Figure 2.**
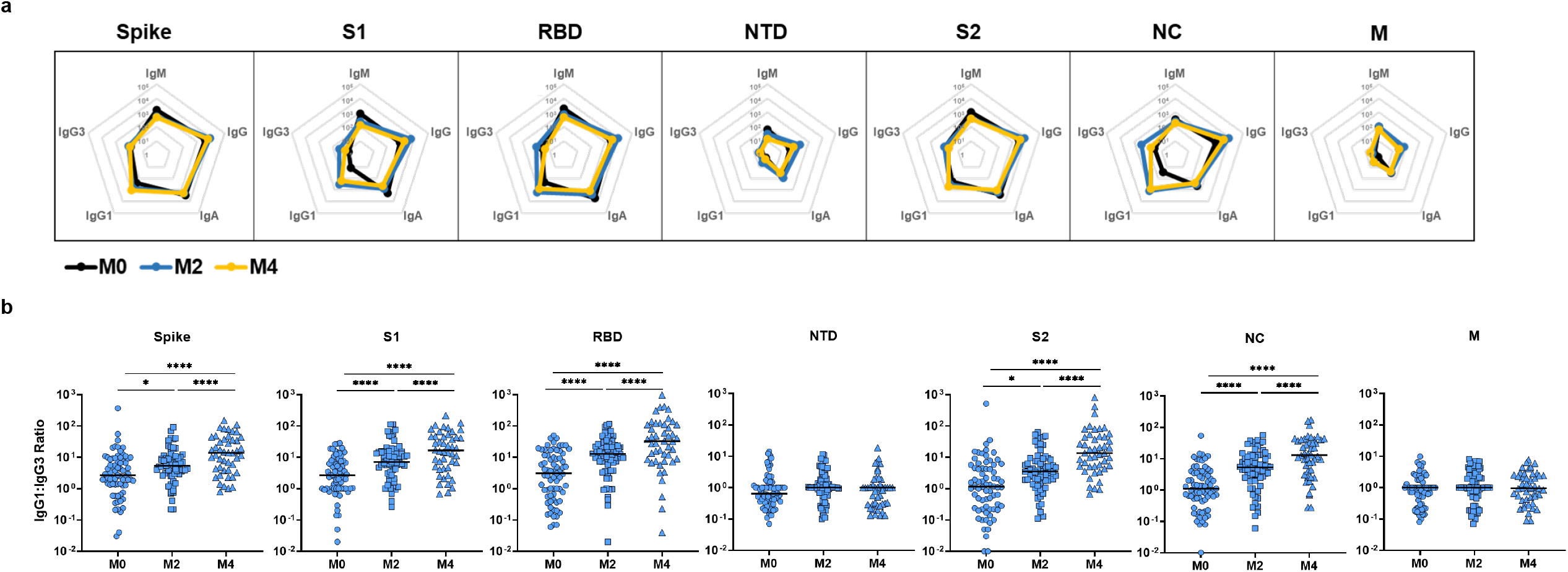
SARS-CoV-2 antigen-specific antibody isotypes and IgG subclass binding. Antigen-specific binding to spike, subunit 1 (S1), receptor binding domain (RBD), N-terminal domain (NTD), subunit 2 (S2), nucleocapsid (NC), and membrane (M) proteins was measured using a Luminex-based multiplex assay for IgM, IgG, IgG1, IgG3, and IgA. The graphs depict (**a**) the distribution of each antibody isotype or subclass represented as mean fluorescence intensity (MFI) and (**b**) the IgG1:IG3 ratio for each of the antigens. Data are shown for binding during the acute phase of infection (M0), 2 months after acute infection (M2), and 4 months after acute infection (M4). IgG1:IgG3 ratios were compared across time points using Wilcoxon signed-rank tests. * p<0.05; ** p<0.01; *** p<0.005; **** p<0.0001

**Supplemental Figure 3.**
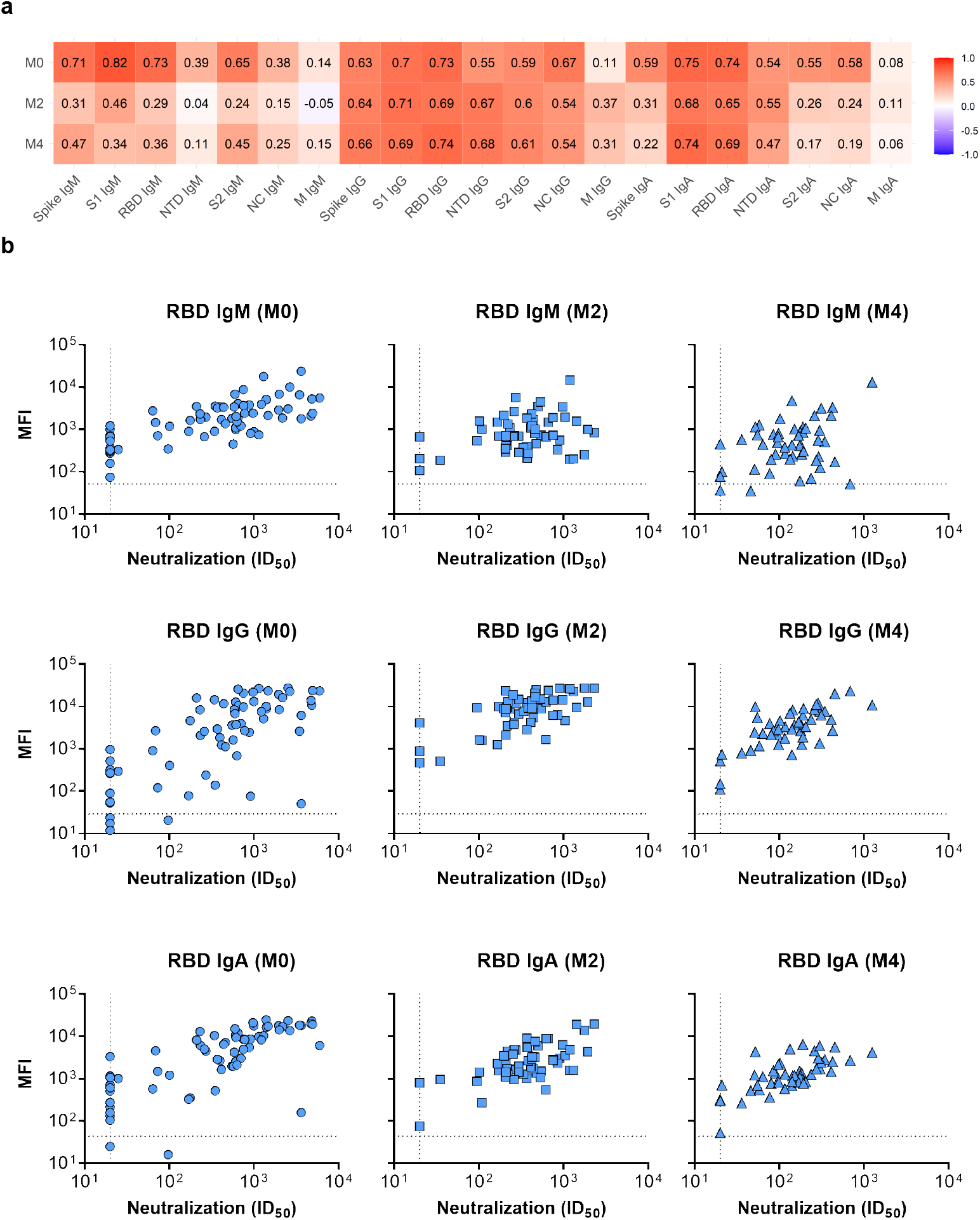
Correlation between levels of SARS-CoV-2-specific antibodies and neutralizing activity (ID_50_) in sera from children and adolescents. Levels of IgM, IgG, and IgA antibodies to SARS-CoV-2 antigens were measured as mean fluorescence intensity using Luminex-based multiplex assays. Antibody-mediated neutralization activity was measured using a pseudovirus (614G) assay and is presented as 50% inhibitory dilution (ID50). (a) Pearson’s correlation coefficients (r) are shown between specific antibody measures and ID50, each considered on the logarithmic scale. (b) Correlation plots between RBD-specific antibodies (IgM, IgG, IgA) and neutralization.

**Supplemental Figure 4.**
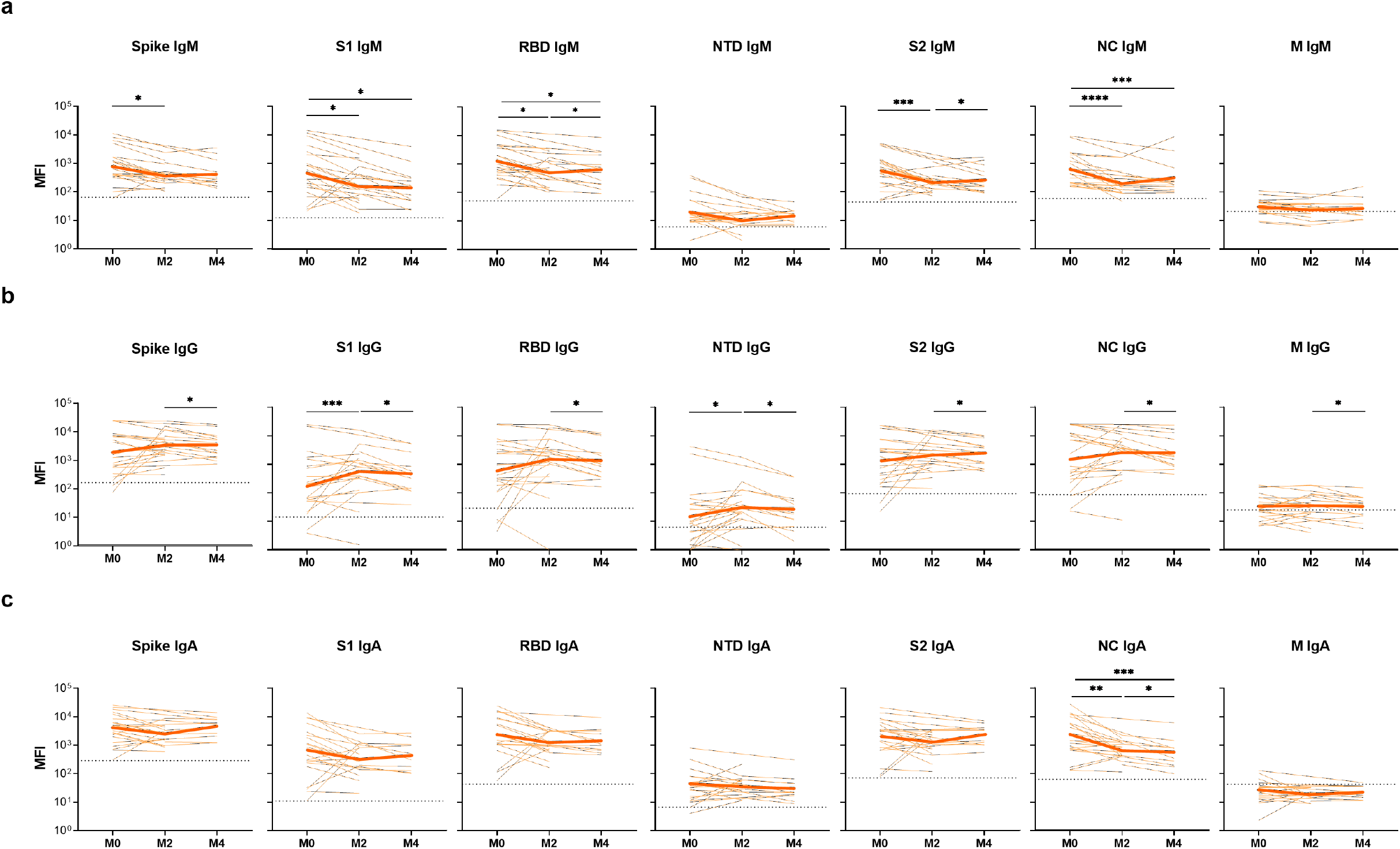
SARS-CoV-2-specific binding antibodies in sera from adults. Specific binding to SARS-CoV-2 antigens (spike, subunit 1 (S1), subunit 2 (S2), receptor binding domain (RBD), N-terminal domain (NTD), subunit 2 (S2), nucleocapsid (NC), and membrane (M) proteins) was measured by a Luminex-based multiplex assay for IgM (**a**), IgG (**b**), and IgA (**c**). Mean fluorescence intensity (MFI) of sera from specific individuals (light orange lines) and the geometric mean in all individuals (thick orange lines) are shown at the time of acute infection (M0) and 2 months (M2) and 4 months (M4) after acute infection. The dotted lines indicate assay thresholds corresponding to the mean MFI plus three standard deviations in sera from 10 SARS-CoV-2-uninfected individuals. Levels of specific antibodies were compared across time points using Wilcoxon signed-ranks tests. * p<0.05; ** p<0.01; *** p<0.005; **** p<0.0001.

**Supplemental Figure 5.**
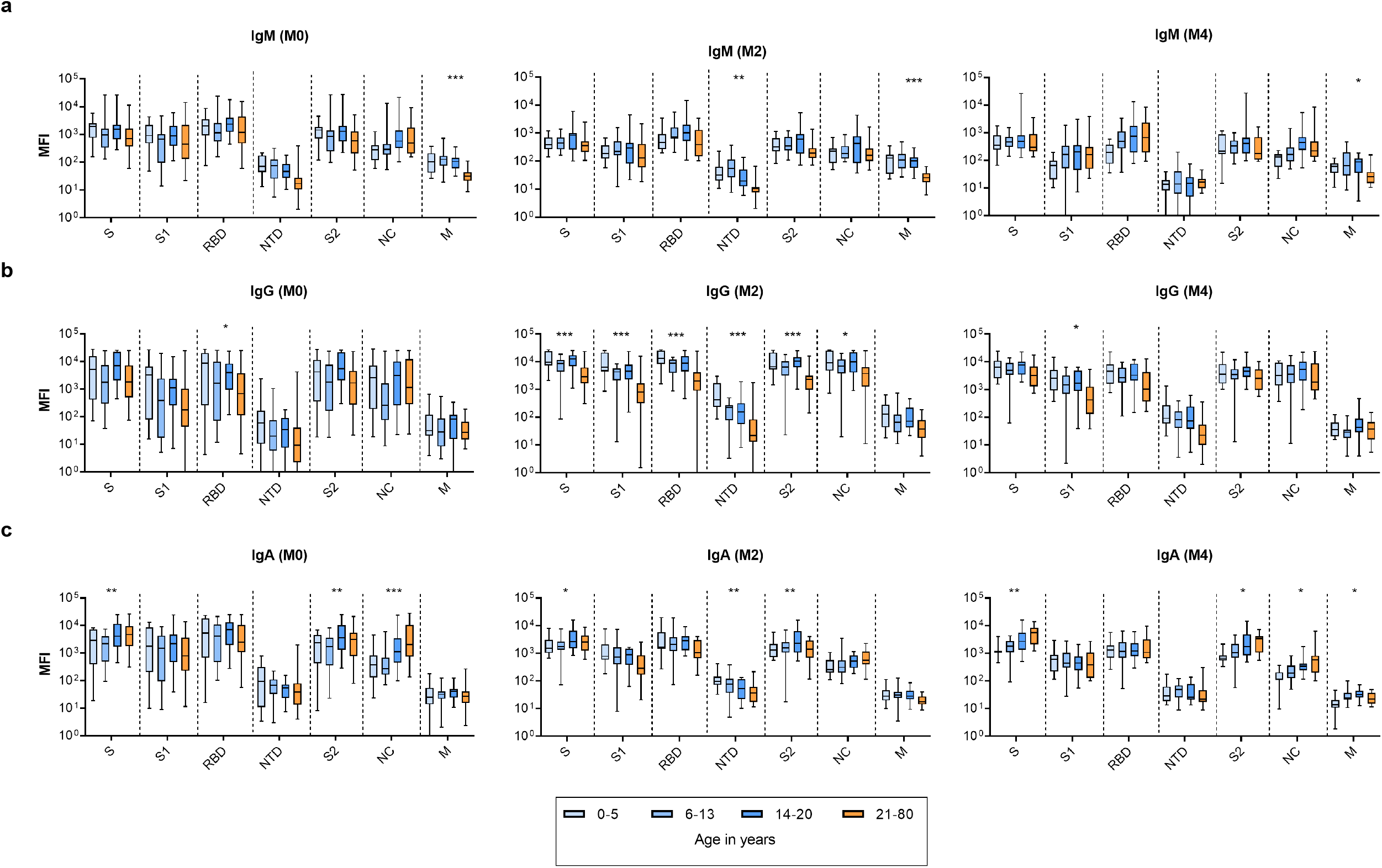
SARS-CoV-2 binding antibodies in sera by age group. Specific binding to SARS-CoV-2 antigens (Spike (S), subunit 1 (S1), subunit 2 (S2), receptor binding domain (RBD), N-terminal domain (NTD), subunit 2 (S2), nucleocapsid (NC), and membrane (M) proteins) was measured by a Luminex-based multiplex assay for IgM (**a**), IgG (**b**), and IgA (**c**). The dotted lines indicate assay thresholds corresponding to the mean MFI plus three standard deviations in sera from 10 SARS-CoV-2-uninfected individuals. Levels of specific antibodies were compared across age category using ANOVA. * p<0.05; ** p<0.01; *** p<0.005; **** p<0.0001

**Supplemental Figure 6.**
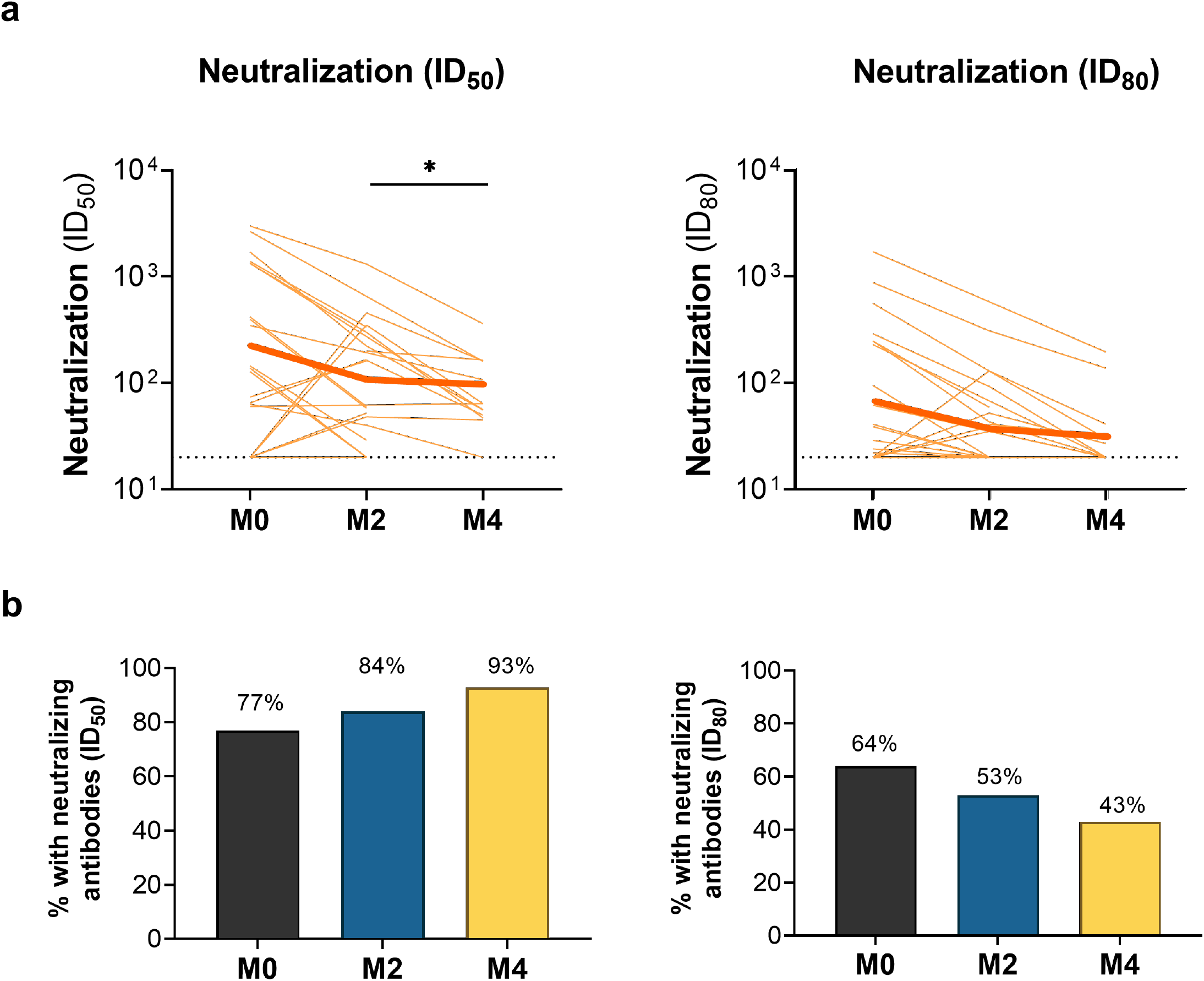
SARS-CoV-2 neutralizing antibodies in sera from adults. Antibody-mediated neutralization activity was measured using a pseudovirus (614G) assay and is presented as 50% inhibitory dilution (ID50) or 80% inhibitory dilution (ID80). (a) Neutralization activity in sera from specific individuals (light orange lines) and the geometric mean in all individuals (thick orange lines) is shown at the time of acute infection (M0) and 2 months (M2) and 4 months (M4) after acute infection. Dotted lines indicate assay positivity thresholds. Comparisons of samples from individuals across time points were made using Wilcoxon signed-rank tests. We did not detect any significant differences across timepoints for either ID50 or ID80. (b) Proportion of adults with detectable neutralizing antibodies at ID50 (left graph) and ID_80_ (right graph) at specific time points after acute infection.

**Table S1.** SARS-CoV-2-specific binding antibodies amongst children and adolescents (0-20 years of age)

|  |  | Acute Infection<br>(M0; n=69) |  | Month 2<br>(M2: n=56) |  | Month 4<br>(M4; n=50) |  |
| --- | --- | --- | --- | --- | --- | --- | --- |
|  |  | Median (IQR) MFI |  | Median (IQR) MFI |  | Median (IQR) MFI |  |
| IgM |  |  |  |  |  |  |  |
|  | Spike | 1434.5 | (658.8, 2047.3) | 506.4 | (266.3, 839.1) | 472.2 | (308.0, 783.3) |
|  | Subunit 1 | 814.5 | (331.8, 1609.5) | 238.9 | (147.9, 442.6) | 127.5 | (46.5, 265.3) |
|  | RBD | 1825.0 | (803.5, 3398.3) | 721.1 | (414.3, 1504.1) | 446.9 | (194.8, 922.7) |
|  | NTD | 65.0 | (28.3, 111.5) | 32.4 | (17.4, 65.1) | 14.2 | (6.9, 25.6) |
|  | Subunit 2 | 1054.3 | (566.5, 1682.3) | 379.2 | (225.7, 725.0) | 358.1 | (172.2, 624.2) |
|  | NC | 330.3 | (193.3, 568.5) | 222.8 | (108.3, 359.5) | 175.1 | (97.0, 429.2) |
|  | Membrane | 105.0 | (61.0, 164.0) | 108.5 | (54.8, 166.7) | 66.0 | (36.1, 114.6) |
| IgG |  |  |  |  |  |  |  |
|  | Spike | 3995.0 | (726.8, 12348.5) | 9221.0 | (4980.0, 15926.0) | 6211.2 | (3513.0, 9483.5) |
|  | Subunit 1 | 876.3 | (77.5, 3709.5) | 5131.0 | (2566.0, 7838.0) | 1751.4 | (836.6, 3375.6) |
|  | RBD | 2912.8 | (294.8, 10687.0) | 9360.0 | (4520.0, 14450.5) | 3494.3 | (1941.4, 6721.1) |
|  | NTD | 41.5 | (6.3, 92.5) | 231.4 | (113.7, 347.1) | 77.5 | (49.7, 217.0) |
|  | Subunit 2 | 3120.0 | (606.5, 11443.5) | 8272.0 | (4680.0, 13220.0) | 3642.0 | (2828.0, 6040.0) |
|  | NC | 962.8 | (134.0, 4370.0) | 8554.0 | (4475.0, 15361.2) | 3558.6 | (1763.6, 7694.8) |
|  | Membrane | 30.8 | (15.8, 107.5) | 73.7 | (38.2, 160.6) | 31.8 | (21.0, 57.3) |
| IgA |  |  |  |  |  |  |  |
|  | Spike | 2834.5 | (884.8, 6293.0) | 1823.9 | (1274.6, 3517.1) | 1681.5 | (1085.2, 3061.2) |
|  | Subunit 1 | 1750.5 | (347.5, 4579.0) | 763.8 | (528.1, 1517.7) | 450.2 | (266.4, 805.7) |
|  | RBD | 5006.0 | (1060.0, 11851.0) | 2267.0 | (1403.0, 4331.0) | 1204.0 | (786.0, 2256.0) |
|  | NTD | 66.8 | (23.3, 117.3) | 81.3 | (39.6, 118.5) | 32.8 | (22.5, 64.7) |
|  | Subunit 2 | 2462.0 | (548.0, 4450.0) | 1510.9 | (969.9, 2902.6) | 1044.8 | (613.4, 2108.7) |
|  | NC | 434.3 | (216.3, 1109.8) | 331.6 | (231.7, 605.4) | 245.1 | (128.5, 360.0) |
|  | Membrane | 31.8 | (19.0, 45.0) | 28.8 | (21.2, 41.6) | 24.4 | (14.9, 36.2) |

IQR, interquartile range; MFI, mean fluorescence intensity; RBD, receptor binding domain; NTD, N-terminal domain; NC, nucleocapsid

**Table S2.**
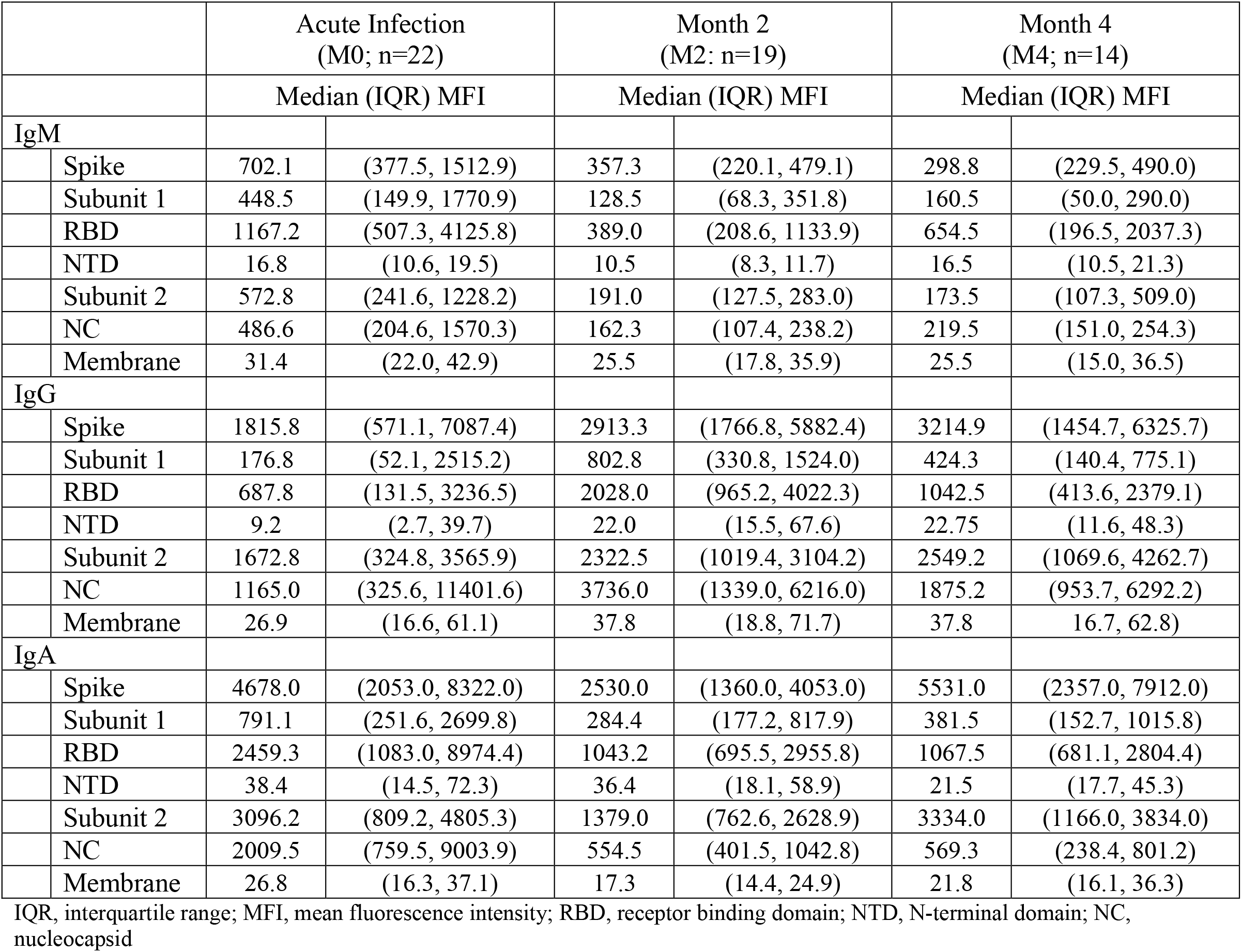
SARS-CoV-2-specific binding antibodies amongst adults (21-70 years of age)

## REFERENCES

1. Johns Hopkins University Coronavirus Resource Center. Accessed February 14, 2021. https://coronavirus.jhu.edu/data

2. Wajnberg A, Amanat F, Firpo A, et al. Robust neutralizing antibodies to SARS-CoV-2 infection persist for months. Science. 2020;370(6521):1227–1230. doi:10.1126/science.abd7728

3. Dan JM, Mateus J, Kato Y, et al. Immunological memory to SARS-CoV-2 assessed for up to 8 months after infection. Science. 2021;371(6529):eabf4063. doi:10.1126/science.abf4063

4. Long Q-X, Tang X-J, Shi Q-L, et al. Clinical and immunological assessment of asymptomatic SARS-CoV-2 infections. Nat Med. 2020;26(8):1200–1204. doi:10.1038/s41591-020-0965-6

5. Kong W-H, Zhao R, Zhou J-B, et al. Serologic Response to SARS-CoV-2 in COVID-19 Patients with Different Severity. Virol Sin. 2020;35(6):752–757. doi:10.1007/s12250-020-00270-x

6. Hurst JH, Heston SM, Chambers HN, et al. SARS-CoV-2 Infections Among Children in the Biospecimens from Respiratory Virus-Exposed Kids (BRAVE Kids) Study. Clin Infect Dis. Published online November 3, 2020. doi:10.1093/cid/ciaa1693

7. Singh T, Heston SM, Langel SN, et al. Lessons From COVID-19 in Children: Key Hypotheses to Guide Preventative and Therapeutic Strategies. Clin Infect Dis. 2020;71(8):2006–2013. doi:10.1093/cid/ciaa547

8. Viner RM, Ward JL, Hudson LD, et al. Systematic review of reviews of symptoms and signs of COVID-19 in children and adolescents. Arch Dis Child. Published online December 17, 2020. doi:10.1136/archdischild-2020-320972

9. Laws RL, Chancey RJ, Rabold EM, et al. Symptoms and Transmission of SARS-CoV-2 Among Children — Utah and Wisconsin, March–May 2020. Pediatrics. 2021;147(1):e2020027268. doi:10.1542/peds.2020-027268

10. Han MS, Choi EH, Chang SH, et al. Clinical Characteristics and Viral RNA Detection in Children With Coronavirus Disease 2019 in the Republic of Korea. JAMA Pediatr. Published online August 28, 2020. doi:10.1001/jamapediatrics.2020.3988

11. Prendergast AJ, Klenerman P, Goulder PJR. The impact of differential antiviral immunity in children and adults. Nat Rev Immunol. 2012;12(9):636–648. doi:10.1038/nri3277

12. Channappanavar R, Perlman S. Age-related susceptibility to coronavirus infections: role of impaired and dysregulated host immunity. J Clin Invest. 2020;130(12):6204–6213. doi:10.1172/JCI144115

13. Allen JC, Toapanta FR, Chen W, Tennant SM. Understanding immunosenescence and its impact on vaccination of older adults. Vaccine. 2020;38(52):8264–8272. doi:10.1016/j.vaccine.2020.11.002

14. Panda A, Qian F, Mohanty S, et al. Age-associated decrease in TLR function in primary human dendritic cells predicts influenza vaccine response. J Immunol. 2010;184(5):2518–2527. doi:10.4049/jimmunol.0901022

15. Archer B, Cohen C, Naidoo D, et al. Interim report on pandemic H1N1 influenza virus infections in South Africa, April to October 2009: epidemiology and factors associated with fatal cases. Euro Surveill. 2009;14(42). doi:10.2807/ese.14.42.19369-en

16. Cao W, Jamieson BD, Hultin LE, Hultin PM, Effros RB, Detels R. Premature aging of T cells is associated with faster HIV-1 disease progression. J Acquir Immune Defic Syndr. 2009;50(2):137–147. doi:10.1097/QAI.0b013e3181926c28

17. Yan Z, Maecker HT, Brodin P, et al. Aging and CMV discordance are associated with increased immune diversity between monozygotic twins. Immun Ageing. 2021;18(1):5. doi:10.1186/s12979-021-00216-1

18. Weisberg SP, Connors TJ, Zhu Y, et al. Distinct antibody responses to SARS-CoV-2 in children and adults across the COVID-19 clinical spectrum. Nat Immunol. 2021;22(1):25–31. doi:10.1038/s41590-020-00826-9

19. Feldstein LR, Rose EB, Horwitz SM, et al. Multisystem Inflammatory Syndrome in U.S. Children and Adolescents. N Engl J Med. 2020;383(4):334–346. doi:10.1056/NEJMoa2021680

20. Post N, Eddy D, Huntley C, et al. Antibody response to SARS-CoV-2 infection in humans: A systematic review. PLoS One. 2020;15(12):e0244126. doi:10.1371/journal.pone.0244126

21. Pierce CA, Preston-Hurlburt P, Dai Y, et al. Immune responses to SARS-CoV-2 infection in hospitalized pediatric and adult patients. Sci Transl Med. 2020;12(564):eabd5487. doi:10.1126/scitranslmed.abd5487

22. Rostad CA, Chahroudi A, Mantus G, et al. Quantitative SARS-CoV-2 Serology in Children With Multisystem Inflammatory Syndrome (MIS-C). Pediatrics. 2020;146(6). doi:10.1542/peds.2020-018242

23. Korber B, Fischer WM, Gnanakaran S, et al. Tracking Changes in SARS-CoV-2 Spike: Evidence that D614G Increases Infectivity of the COVID-19 Virus. Cell. 2020;182(4):812-827.e19. doi:10.1016/j.cell.2020.06.043

24. Lau EHY, Tsang OTY, Hui DSC, et al. Neutralizing antibody titres in SARS-CoV-2 infections. Nat Commun. 2021;12(1):63. doi:10.1038/s41467-020-20247-4

25. Dogan M, Kozhaya L, Placek L, et al. SARS-CoV-2 specific antibody and neutralization assays reveal the wide range of the humoral immune response to virus. Commun Biol. 2021;4(1):129. doi:10.1038/s42003-021-01649-6

26. Carsetti R, Zaffina S, Piano Mortari E, et al. Different Innate and Adaptive Immune Responses to SARS-CoV-2 Infection of Asymptomatic, Mild, and Severe Cases. Front Immunol. 2020;11:610300. doi:10.3389/fimmu.2020.610300

27. the UNICORN Consortium, Milani GP, Dioni L, et al. Serological follow-up of SARS-CoV-2 asymptomatic subjects. Sci Rep. 2020;10(1):20048. doi:10.1038/s41598-020-77125-8

28. Jiang C, Wang Y, Hu M, et al. Antibody seroconversion in asymptomatic and symptomatic patients infected with severe acute respiratory syndrome coronavirus 2 (SARS-CoV-2). Clin Transl Immunol. 2020;9(9). doi:10.1002/cti2.1182

29. Ko J-H, Joo E-J, Park S-J, et al. Neutralizing Antibody Production in Asymptomatic and Mild COVID-19 Patients, in Comparison with Pneumonic COVID-19 Patients. JCM. 2020;9(7):2268. doi:10.3390/jcm9072268

30. Amanat F, Stadlbauer D, Strohmeier S, et al. A serological assay to detect SARS-CoV-2 seroconversion in humans. Nat Med. 2020;26(7):1033–1036. doi:10.1038/s41591-020-0913-5

31. Long Q-X, Liu B-Z, Deng H-J, et al. Antibody responses to SARS-CoV-2 in patients with COVID-19. Nat Med. 2020;26(6):845–848. doi:10.1038/s41591-020-0897-1

32. Luchsinger LL, Ransegnola BP, Jin DK, et al. Serological Assays Estimate Highly Variable SARS-CoV-2 Neutralizing Antibody Activity in Recovered COVID-19 Patients. Loeffelholz MJ, ed. J Clin Microbiol. 2020;58(12):e02005–20, /jcm/58/12/JCM.02005-20.atom. doi:10.1128/JCM.02005-20

33. Okba NMA, Müller MA, Li W, et al. Severe Acute Respiratory Syndrome Coronavirus 2-Specific Antibody Responses in Coronavirus Disease Patients. Emerg Infect Dis. 2020;26(7):1478–1488. doi:10.3201/eid2607.200841

34. Prévost J, Gasser R, Beaudoin-Bussières G, et al. Cross-Sectional Evaluation of Humoral Responses against SARS-CoV-2 Spike. Cell Rep Med. 2020;1(7):100126. doi:10.1016/j.xcrm.2020.100126

35. Gudbjartsson DF, Norddahl GL, Melsted P, et al. Humoral Immune Response to SARS-CoV-2 in Iceland. N Engl J Med. 2020;383(18):1724–1734. doi:10.1056/NEJMoa2026116

36. Plotkin SA. Correlates of Protection Induced by Vaccination. CVI. 2010;17(7):1055–1065. doi:10.1128/CVI.00131-10

37. Hassan AO, Case JB, Winkler ES, et al. A SARS-CoV-2 Infection Model in Mice Demonstrates Protection by Neutralizing Antibodies. Cell. 2020;182(3):744-753.e4. doi:10.1016/j.cell.2020.06.011

38. Iyer AS, Jones FK, Nodoushani A, et al. Persistence and decay of human antibody responses to the receptor binding domain of SARS-CoV-2 spike protein in COVID-19 patients. Sci Immunol. 2020;5(52):eabe0367. doi:10.1126/sciimmunol.abe0367

39. Rodda LB, Netland J, Shehata L, et al. Functional SARS-CoV-2-Specific Immune Memory Persists after Mild COVID-19. Cell. 2021;184(1):169-183.e17. doi:10.1016/j.cell.2020.11.029

40. PFIZER-BIONTECH ANNOUNCE POSITIVE TOPLINE RESULTS OF PIVOTAL COVID-19 VACCINE STUDY IN ADOLESCENTS. Published March 31, 2021. https://www.pfizer.com/news/press-release/press-release-detail/pfizer-biontech-announce-positive-topline-results-pivotal

41. Nie J, Li Q, Wu J, et al. Establishment and validation of a pseudovirus neutralization assay for SARS-CoV-2. Emerging Microbes & Infections. 2020;9(1):680–686. doi:10.1080/22221751.2020.1743767

42. McClain MT, Constantine FJ, Henao R, et al. Dysregulated transcriptional responses to SARS-CoV-2 in the periphery. Nat Commun. 2021;12(1):1079. doi:10.1038/s41467-021-21289-y

43. Benjamini Y, Hochberg Y. Controlling the False Discovery Rate -a Practical and Powerful Approach. J R Stat Soc Ser B-Methodol. 1995;57(1):289–300.

44. Harris PA, Taylor R, Thielke R, Payne J, Gonzalez N, Conde JG. Research electronic data capture (REDCap)--a metadata-driven methodology and workflow process for providing translational research informatics support. J Biomed Inform. 2009;42(2):377–381. doi:10.1016/j.jbi.2008.08.010

45. R Core Team. R: A language and environment for statistical computing. R Foundation for Statistical Computing. http://www.R-project.org/

